# Validation of Patient-Reported Outcomes Measurement Information System^®^ (PROMIS^®^) Pediatric Measures for Children with Chronic Nonbacterial Osteomyelitis

**DOI:** 10.1101/2025.09.13.25335689

**Authors:** Mary M. Eckert, Eveline Y. Wu, Melissa Oliver, Joshua Scheck, Sivia Lapidus, Ummusen Kaya Akca, Shima Yasin, Aleksander Lenert, Sara Stern, Antonella Insalaco, Manuela Pardeo, Gabriele Simonini, Edoardo Marrani, Xing Wang, Bin Huang, Leonard K. Kovalick, Natalie Rosenwasser, Erin Balay-Destrude, Gabriel Casselman, Adriel Liau, Ava Klein, Yurong Shao, Claire Yang, Molly Briggs, Ethan Mueller, Emily Deng, Paige Rhiannon Trunnell, Iris Hamilton, Elise Machrone, Doaa Mosad Mosa, Lori Tucker, Hermann Girschick, Ronald M. Laxer, Georgina Tiller, Jonathan Akikusa, Christian M. Hedrich, Karen Onel, Fatma Dedeoglu, Marinka Twilt, Seza Ozen, Polly J Ferguson, Laura Schanberg, Bryce B. Reeve, Yongdong Zhao

## Abstract

**Objectives:** To assess the validity of the Patient-Reported Outcomes Measurement Information System^®^ (PROMIS^®^) Pediatric measures in patients with chronic nonbacterial osteomyelitis (CNO).

**Methods:** Within the longitudinal patient registry of CNO, English-speaking patients aged 8 years and older self-reported PROMIS Pediatric measures of fatigue, pain interference (PI), pain behavior (PB), mobility, upper extremity (UE), physical activity (PA) and strength impact (SI), and external validation measures. Log-transformed linear mixed-effects models with random patient intercepts were used to assess PROMIS T-score changes. Wilcoxon signed-rank test was performed to determine the PROMIS T-score changes among the improved, worsened, and unchanged groups. Spearman rank correlation test determined the relationship of PROMIS T-scores with disease status reported by patients/families.

**Results:** More than 1,000 clinical visits from 184 patients included PROMIS Pediatric measures entries. All PROMIS T-scores correlated significantly (p<0.01) with patient-reported variables and physician global assessment (PHGA). The correlation between function and mobility, PB, and PI was good (r=0.4-0.6). The correlation of patient-reported disease status was strong with PHGA (r=0.75), moderate with mobility, PB, PI, and weak with Fatigue, SI, UE, and PA. The changes of PROMIS T-scores over time for mobility, PB, PA, and PI compared to the self-reported status change were significant (p<0.05). After effective treatment, when clinical disease activity score improved by at least 2.5 points (n=18), the change of PROMIS T-scores for mobility, PB, PI, UE were significant (p<0.05).

**Conclusion:** This study provided evidence supporting the use of PROMIS Pediatric mobility, pain behavior, and pain interference measures for CNO clinical disease monitoring.

**Key Messages:** PROMIS Pediatric measures are valid tools for assessing disease burden in patients with Chronic Nonbacterial osteomyelitis (CNO); T-scores across all domains showed significant correlations with patient-reported outcomes and physician global assessments, with particularly strong correlations for mobility, pain behavior, and pain interference.

PROMIS Pediatric T-scores for mobility, pain behavior, physical activity, and pain interference were sensitive to patient-reported changes in disease status over time, supporting their use in tracking disease progression in CNO.

Following effective treatment, defined by at least a 2.5-point improvement in clinical disease activity score, PROMIS Pediatric T-scores for mobility, pain behavior, pain interference, and upper extremity function showed significant improvement, highlighting their responsiveness to treatment in CNO.

## Background

Chronic nonbacterial osteomyelitis (CNO), also known as chronic recurrent multifocal osteomyelitis (CRMO), is a rare autoinflammatory bone disease characterized by destruction of the bone, persistent pain, and an increased risk of fractures^1^. CNO significantly impacts quality of life and socioeconomic well-being. ^2^ Families frequently report that CNO interferes with their children’s daily functioning and ability to participate in recreational activities ^3^. Although physical performance tests in some studies did not show major physiological limitations, patient-reported outcomes have consistently demonstrated reduced quality of life^3,4^. In one study using exercise tests with gas exchange and accelerometry, no significant differences were observed in exercise capacity between patients with CNO and healthy controls. ^5^ However, patients with CNO participated in fewer sports and scored lower on most domains of the Pediatric Quality of Life Inventory (PedsQL) 3.0 and 4.0, indicating a diminished health-related quality of life (HRQOL). ^5^ In a separate prospective study, the global patient/parent assessment (0-10 scale) score improved from 3.8 at baseline to 0.5 after 12 months of naproxen treatment. ^6^ The Childhood Health Assessment Questionnaire (CHAQ) score similarly improved from 0.7 to 0, reflecting responsiveness to treatment. ^6^ These findings highlight the importance of capturing the patient and family experience, including daily functioning, participation, and overall quality of life, as an essential component of disease assessment. The Patient-Reported Outcomes Measurement Information System^®^ (PROMIS^®^) was developed to provide a standardized, scalable approach to assessing key HRQOL domains, including pain, fatigue, physical function, and emotional well-being, across a variety of conditions. PROMIS Pediatric measures have shown value in pediatric rheumatic diseases such as juvenile idiopathic arthritis (JIA) and systemic lupus erythematosus (SLE), particularly in distinguishing active from inactive disease and assessing treatment response. ^7^ However, no studies to date have evaluated PROMIS Pediatric measures for patients with CNO. Given that CNO differs in both pathophysiology and symptom pattern from other rheumatic diseases, such as JIA and SLE, additional evidence for the validity of PROMIS Pediatric measures are warranted using a well-curated cohort of patients. To address this gap, we evaluated PROMIS Pediatric measures in a prospective, longitudinal, international cohort study: the Chronic Nonbacterial Osteomyelitis International Registry (CHOIR).^8^

## Patients and Methods

### Patient cohort

Patients diagnosed with CNO and under 18 years of age at the time of enrollment were prospectively enrolled in the Chronic Nonbacterial Osteomyelitis International Registry (CHIOR) (Seattle Children’s Hospital IRB#1232). The CHIOR study is an international observational longitudinal cohort study of patients with CNO established by the Childhood Arthritis and Rheumatology Research Alliance (CARRA) CNO workgroup. All participating sites provided standard of clinical care for patient enrollment and used standardized outcome measurements. Enrolment occurred across 14 sites (8 U.S. sites, 2 Canadian sites, 2 Italian sites, 1 Egyptian site, 1 Turkish site) starting August 1, 2018. Informed consent was obtained for all patients and, additionally, assent was obtained from age-appropriate patients. We excluded patients from the international sites whose official languages do not include English. Institutional Board Review approval was obtained at each registry site. Between August 1, 2018, and September 30, 2021, enrollment was limited to patients with NSAID-refractory disease (defined as no improvement after 1-3 months of NSAID therapy) and/or active spinal lesions as confirmed by MRI. After October 1, 2021, enrollment criteria expanded to include all patients with CNO regardless of disease severity or treatment status.

### Administration of PROMIS and validation questionnaires

PROMIS tools were developed through NIH initiative to measure pain, fatigue, physical functioning, emotional distress, and social role participation that have a major impact on HRQOL across a variety of chronic diseases^9,10^. PROMIS Pediatric measures in static short forms and CHAQ, a commonly used assessment in pediatric rheumatology, were administered at clinic visits (in person or virtual) via hardcopy or electronic forms^11^. Patients were included if they had a visit date before January 2023 and aged ≥ 8 years at the time of the visit as well as completing at least one PROMIS questionnaire in English. PROMIS Pediatric domains included fatigue, pain interference (PI), pain behavior (PB), mobility, upper extremity function (UE), physical activity (PA), and strength impact (SI). Higher PROMIS T-scores for symptoms (fatigue, pain interference, pain behavior) reflect worse symptom burden, and higher PROMIS T-scores for function (mobility, UE, PA, SI) reflect better functioning. Patients also completed other *questionnaires* that included questions measuring degrees of difficulty using arms/legs/trunk/jaw, fatigue, depression, worry/anxiety, pain, and patient/parent global assessment of disease activity using a 0-10 numeric rating scale through self-reporting. Additional questions (**supplement 1**) assessing self-perceived disease activity (inactive, mildly active, moderately active, or severely active), changes in disease activity compared to the previous visit (improved, worsened, or unchanged), and perceptions of the treatment effectiveness were also administered to patients.

### Clinical data collection

Clinical assessments included 1) patient pain assessment scored on a 0-10 Likert scale, 2) patient/parent global assessment of disease activity scored on a 0-10 Likert scale, 3) physician global assessment of disease activity (PHGA) scored on a 0-10 Likert scale, and 4) number of clinically active CNO lesions defined as focal tenderness, and/or swelling, and/or warmth in addition to patient’s report of pain at a known CNO lesion site. Physicians were blinded to the patient’s questionnaire responses when completing the PHGA scores. A CNO clinical disease activity score (CDAS) was calculated by summing the patient pain and patient/parent global assessment of disease activity scores and total number of clinically active CNO lesions. Improvement of CDAS of ≥2.5 was defined as clinically meaningful change^8^.

### Statistical analyses

Descriptive statistics were used to summarize demographic and clinical characteristics. PROMIS Pediatric T-score changes after treatment were evaluated using log-transformed linear mixed effect models with random patient intercepts. The Wilcoxon signed-rank test with continuity correction was performed to determine changes in PROMIS T-scores among the patient-reported improved, worsened, and unchanged groups. Spearman rank correlation test was used to assess the relationship between PROMIS T-scores and reported disease status by patient/families^12,13^. Correlations were classified as: small (0.10–0.29), moderate (0.30–0.49), strong (0.50–0.69), and very strong (>0.70), All computations were performed using R software (version 3.6.1, R Foundation for Statistical Computing, Vienna, Austria). All tests were two-sided, and p-values <0.05 were considered significant. Bonferroni correction was also performed for multiple pair comparisons and reported separately.

## Results

### Demographic and Clinical Data

We identified 184 of 290 patients who were enrolled in CHOIR between August 1, 2018 and January 31, 2023, and completed PROMIS and CHAQ questionnaires. These patients have had at least 1 visit with completion of PROMIS/CHAQ and external validation scores. Of the 184 patients, 38% were male, and 83% identified as white. The patients’ average age of onset was 9.4 years, average age of diagnosis 10.4 years, and average age of enrollment 11.4 years. These demographics are consistent with other studies conducted in CNO patient populations (**Table 1**). ^6^

**Table 1.** Demographics of patients with Chronic Nonbacterial Osteomyelitis (n=184)

|  | **Median [IQR], N (%)** |
| --- | --- |
| **Age at Onset** | 9.4 [7.5, 11.3] |
| **Age at Enrollment** | 11.4 [9.5, 13.8] |
| **Age at Diagnosis** | 10.4 [8.4, 12.3] |
| **Sex, male (%)** | 70 (38) |
| **Race # (%)** |  |
| White (Caucasian) | 153 (83) |
| Black, African American, African, or Afro-Caribbean | 10 (5) |
| Asian | 9 (5) |
| Hispanic, Latino, or Spanish origin | 10 (5) |
| Middle Eastern or North African | 2 (1) |
| Native American, American Indian, or Alaskan Native | 2 (1) |
| Native Hawaiian, or Pacific Islander | 3 (2) |
| Prefer not to answer | 4 (2) |
| Other | 4 (2) |

### Association of PROMIS Pediatric Measures’ T-scores with Established Health Measures’ Scores

Of the 184 patients enrolled in the study, over 1000 visits were conducted during which PROMIS Pediatric measures were collected in conjunction with the PHGA (n=1,013-1,071). All the PROMIS measures’ T-scores showed significant correlations (p<0.01) with PHGA (**Table 2**). In addition, over 300 visits (n=311-317) included the collection of PROMIS Pediatric measures along with self-reported patient outcomes of difficulty of using arms or legs or back or jaw, fatigue, sadness and worry. The T-scores of the PROMIS measures of fatigue, mobility, pain behavior, and pain interference all had significant correlations with p-values of <0.01. The remaining PROMIS measures of PA, SI and UE also show significant correlation, albeit at a lower threshold of p <0.05. The PROMIS Pediatric measures of fatigue, pain behavior and pain interference all showed a positive correlation with PHGA and patient self-reported outcomes. The PROMIS variables of mobility, PA, SI and UE all showed negative correlations with the PHGA and the patient self-reported outcomes. Physical Activity, Strength Impact and Upper Extremity showed small correlations (r <|0.3|) and fatigue, mobility, pain behavior and pain interference showed moderate to strong correlations (|0.3|<r<|0.75|).

**Table 2.**
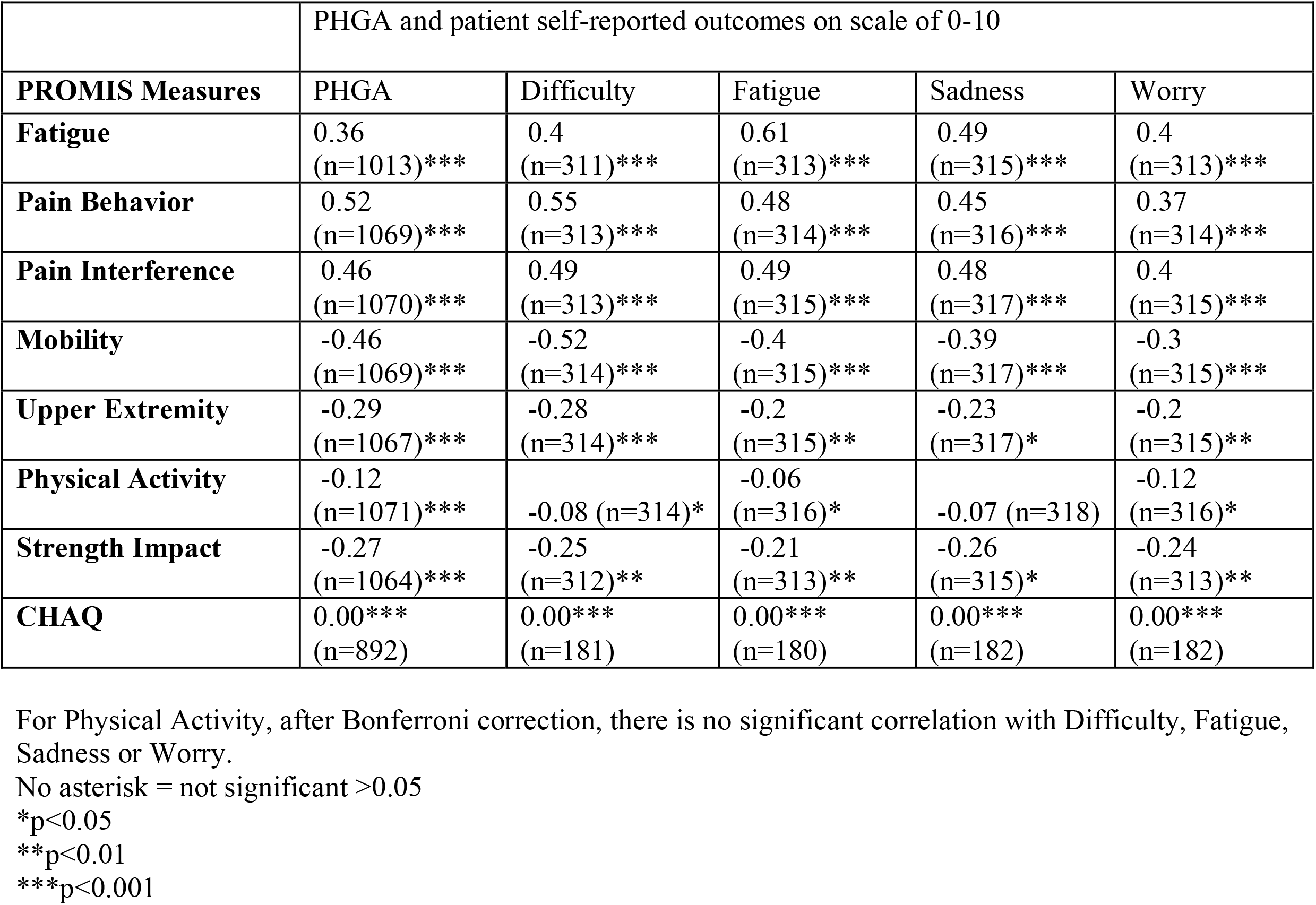
PROMIS Pediatric and CHAQ Scores Correlations with Physician and Patient/Family Reported Outcomes.

|  | PHGA and patient self-reported outcomes on scale of 0-10 |  |  |  |  |
| --- | --- | --- | --- | --- | --- |
| **PROMIS Measures** | PHGA | Difficulty | Fatigue | Sadness | Worry |
| **Fatigue** | 0.36<br>(n=1013)*** | 0.4<br>(n=311)*** | 0.61<br>(n=313)*** | 0.49<br>(n=315)*** | 0.4<br>(n=313)*** |
| **Pain Behavior** | 0.52<br>(n=1069)*** | 0.55<br>(n=313)*** | 0.48<br>(n=314)*** | 0.45<br>(n=316)*** | 0.37<br>(n=314)*** |
| **Pain Interference** | 0.46<br>(n=1070)*** | 0.49<br>(n=313)*** | 0.49<br>(n=315)*** | 0.48<br>(n=317)*** | 0.4<br>(n=315)*** |
| **Mobility** | -0.46<br>(n=1069)*** | -0.52<br>(n=314)*** | -0.4<br>(n=315)*** | -0.39<br>(n=317)*** | -0.3<br>(n=315)*** |
| **Upper Extremity** | -0.29<br>(n=1067)*** | -0.28<br>(n=314)*** | -0.2<br>(n=315)** | -0.23<br>(n=317)* | -0.2<br>(n=315)** |
| **Physical Activity** | -0.12<br>(n=1071)*** | -0.08 (n=314)* | -0.06<br>(n=316)* | -0.07 (n=318) | -0.12<br>(n=316)* |
| **Strength Impact** | -0.27<br>(n=1064)*** | -0.25<br>(n=312)** | -0.21<br>(n=313)** | -0.26<br>(n=315)* | -0.24<br>(n=313)** |
| **CHAQ** | 0.00***<br>(n=892) | 0.00***<br>(n=181) | 0.00***<br>(n=180) | 0.00***<br>(n=182) | 0.00***<br>(n=182) |

For Physical Activity, after Bonferroni correction, there is no significant correlation with Difficulty, Fatigue, Sadness or Worry.
No asterisk = not significant >0.05
\*p<0.05
\*\*p<0.01
\*\*\*p<0.001

### Association of PROMIS Pediatric T-scores with Disease Activity

The PROMIS Pediatric T-scores were also compared with patient-reported disease activity. All PROMIS T-scores except physical activity, correlated significantly (p<0.01) with patient-reported disease activity (**Figure 1**). A very strong correlation was observed between patient-reported disease status and PHGA (r=0.75). Strong correlations of disease status were found with the PROMIS Pediatric measures of Mobility (r=-0.53), pain behavior (r=0.57), and pain interference (r=0.5) and a moderate association with CHAQ (r=0.46). Moderate correlations were observed for fatigue (r=0.36), and small correlations for SI (r=-0.23), and UE (r=-0.23). No significant correlation was found with Physical Activity T-scores (r=-0.03). We then assessed whether changes in PROMIS T-scores were associated with changes in patient-reported disease activity, categorized as self-reported unchanged, improved or worsened status from prior visit. The variables of mobility (p=0.02), pain behavior (p<0.001), physical activity (p=0.02), and pain interference (p=0.01) significantly correlated with the patient-reported disease status. However, PROMIS T-score changes for fatigue (p=0.06), SI (p=0.9), and UE (p=0.09) did not differ across various self-reported status change groups whereas other PROMIS Pediatric T-score changes and CHAQ differ significantly (p<0.05).

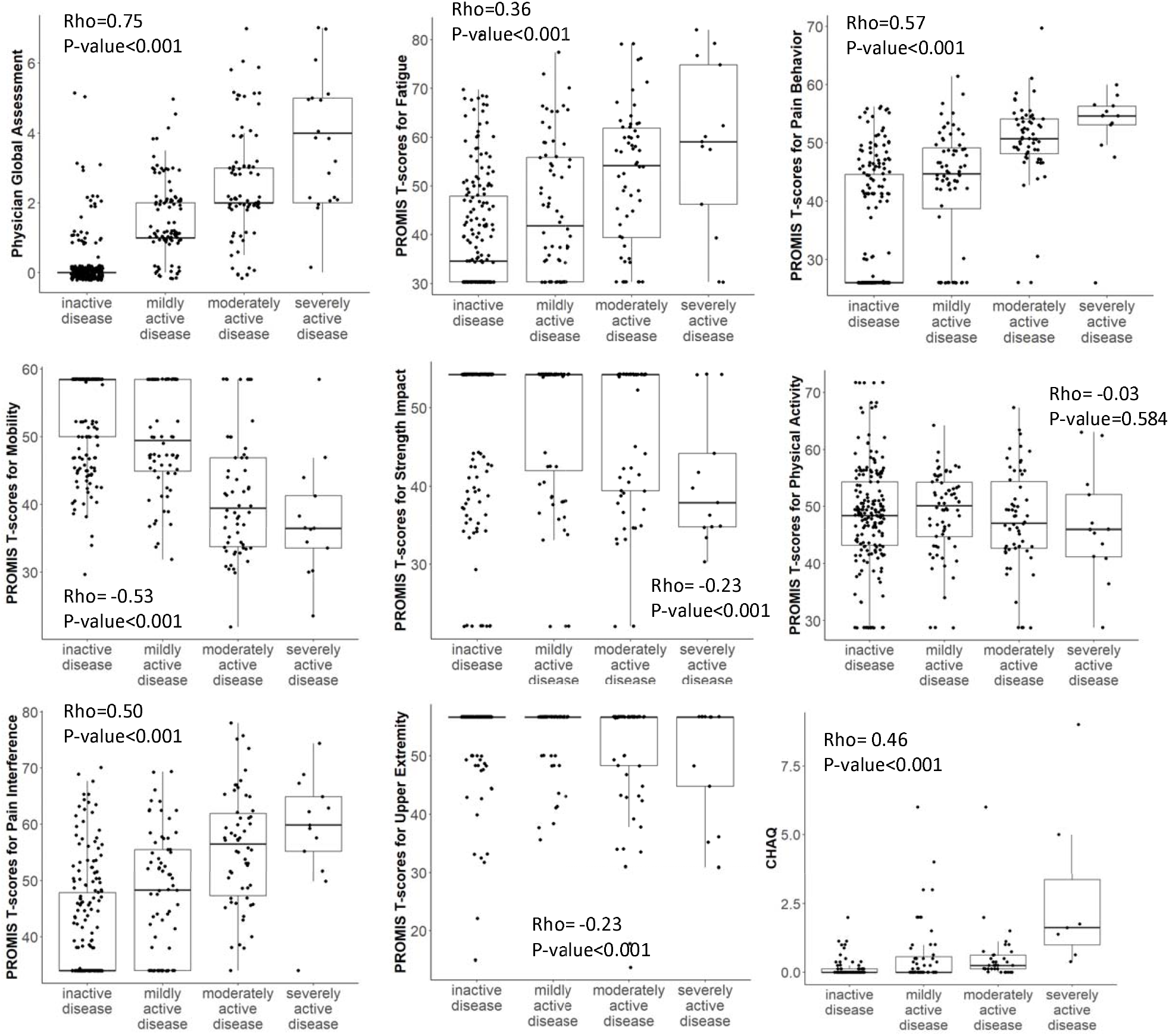

### PROMIS Pediatric T-Score Change Associated with Reported Disease Activity Change

We then compared how the PROMIS Pediatric T-scores changed across time points when patients reported better, unchanged, or worse disease status (**Table 3**). All the PROMIS T-score changes were significant (p <0.05) except fatigue, strength impact, and upper extremity. There is corresponding significant improvement of T-scores of pain behavior, pain interference, mobility and physical activity as well as CHAQ comparing patients with reported “better” than “worse”. The coefficient of variation of these scores were all greater than 100%.

**Table 3.** PROMIS Pediatric Measures T-score or CHAQ score change when patients reported better, unchanged, or worse during follow-up visits.

|  | Score change in Mean (SD) |  |  |  |  |
| --- | --- | --- | --- | --- | --- |
|  | Better | Unchanged | Worse | P-value | Adjusted p value (update) |
| Sample size | n= 47-79 | n= 38-92 | n= 9-29 |  |  |
| Fatigue | -1.9 (11.5) | -0.2 (10.9) | 4.3 (13.2) | 0.06 | 0.08 |
| Pain Behavior | -4.0 (11.3) | -1.9 (8.9) | 9.8 (15.5) | <0.001 | <0.001 |
| Pain Interference | -0.5 (10.5) | -1.5 (7.5) | 4.7 (11.7) | 0.01 | 0.04 |
| Mobility | 0.8 (7.4) | 1.7 (6.1) | -3.1 (11.4) | 0.02 | 0.04 |
| Upper Extremity | 1.1 (5.3) | 1.4 (5.0) | -1 (4.3) | 0.09 | 0.10 |
| Physical Activity | 1.8 (9.3) | 1.5 (9.4) | -3.7 (9.9) | 0.02 | 0.04 |
| Strength Impact | 1.9 (11.2) | 1.2 (11.6) | 0.8 (8.2) | 0.88 | 0.88 |
| CHAQ | -0.2 (0.7) | 0 (0.02) | 0.2 (0.7) | 0.03 | 0.05 |

### The Change of PROMIS T-Scores after introducing Second Line Treatment

A selected group of patients filled out the PROMIS Pediatric measures before and after second line treatments (conventional synthetic or biologic disease modifying anti rheumatic drugs, bisphosphonates) were introduced, and we analyzed the median change from baseline of the PROMIS T-scores, CDAS and CHAQ scores (**Table 4**). All variables were found to be significant except fatigue, physical activity, and strength impact. Second line treatment significantly improved disease activity (CDAS and PHGA), CHAQ, mobility, pain behavior and pain interference. CDAS and PHGA scores showed marked reductions with a median change of -5 [-7, -1] and –2 [-2, 0] respectively. CHAQ also showed score improvement with a median score of 0.25 [0.78, 0]. Among the significant PROMIS Pediatric measures, mobility showed improvement at a median score increase of 2 [0, 11] while pain behavior and pain interference decreased, -3 [-13, 0], and -4 [-15, 0] respectively. In contrast the variables that were found to not be significant, fatigue, physical activity, and strength impact also did not have a large median change in T-score after second line treatment, 0 [-8, 1], -1[-7, 6], and 0 [0, 7] respectively.

**Table 4.** PROMIS Pediatric, PHGA, and CHAQ score changes after second-line treatment.

| PRO measure (n) | Median [IQR] of T score or CHAQ | P value | Adjusted p value (update) |
| --- | --- | --- | --- |
| CDAS (n=22) | -5 [-7, -1] | <0.001 | <0.001 |
| PHGA (n=36) | -2 [-2, 0] | <0.001 | <0.001 |
| Fatigue (N=37) | 0[-8, 1] | 0.27 | 0.30 |
| Pain Behavior (N=37) | -3[-13, 0] | 0.001 | 0.002 |
| Pain Interference (N=37) | -4[-15, 0] | 0.01 | 0.01 |
| Mobility (N=37) | 2[0, 11] | 0.001 | 0.002 |
| Upper Extremity (N=37) | 0[0, 12] | 0.01 | 0.01 |
| Physical Activity (N=37) | -1[-7, 6] | 0.87 | 0.87 |
| Strength Impact (N=37) | 0[0, 7] | 0.15 | 0.19 |
| CHAQ (n= 24) | -0.3[-0.8, 0] | 0.003 | 0.01 |

### Change in PROMIS T-Scores from pre-to post-Effective Treatment

The PROMIS Pediatric measures were evaluated at multiple time points before and after implementation of effective treatment in this patient population (**Table 5**). Specifically, changes in PROMIS T-scores were analyzed in patients who demonstrated a clinically meaningful improvement in disease activity, defined as a ≥ 2.5-point reduction in the Clinical Disease Activity Score (CDAS). Among the PROMIS Pediatric measures, mobility, pain behavior, pain interference, and upper extremity function showed significant improvements (p < 0.05) following effective treatment. In contrast, fatigue, strength impact, and physical activity did not demonstrate statistically significant changes. When comparing paired visits based on the degree of CDAS improvement, patients who exhibited a greater improvement as indicated by a CDAS reduction of at least 2.5, showed significantly greater improvements in PHGA, CHAQ and pain interference scores compared to those with a lesser improvement as indicated by CDAS.

**Table 5.** PROMIS, PHGA, CHAQ score changes between groups with or without CDAS improvement (regardless of what medications)

| PRO Measure | Mean (SD) change of T scores or CHAQ, PHGA when the change of CDAS is >-2.5 (N=8) | Mean (SD) change of T scores or CHAQ, PHGA when the change of CDAS is <-2.5 (N=18) | P value | Adjusted p value (update) |
| --- | --- | --- | --- | --- |
| CDAS | 0 (2) | -7 (3) | **<0.001** | **<0.001** |
| PHGA | 0 (2) | -2 (2) | **0.02** | **0.07** |
| Fatigue | 3 (12) | -6 (16) | 0.15 | 0.21 |
| Pain Behavior | -6 (9) | -6.1 (8) | 0.95 | 0.95 |
| Pain Interference | 3 (10) | -8 (11) | **0.03** | **0.08** |
| Mobility | 2 (7) | 8 (9) | 0.07 | 0.14 |
| Upper Extremity | 0 (0) | 5 (10) | 0.14 | 0.21 |
| Physical Activity | 1 (8) | 2 (10) | 0.86 | 0.95 |
| Strength Impact | 3 (6) | 1 (11) | 0.57 | 0.71 |
| CHAQ | 0.1 (0.2) | -0.4 (0.4) | 0.02 | 0.07 |

## Discussion

Assessing disease outcome in CNO presents a significant challenge due to clinical heterogeneity and the lack of validated, standardized outcome measures. Magnetic resonance imaging (MRI) is frequently used to support diagnosis and monitor disease over time, but its utility is limited by the presence of asymptomatic lesions that can occur earlier or emerge later in the disease course unless it is located in spine. ^14^ Practical barriers to serial imaging, including limited access, high cost and the need for sedation in young children, further restrict MRI’s feasibility for routine monitoring.^15^ Clinical exam and patient pain complaints are not reliable to determine the disease accurately by themselves either. These limitations underscore the need for a more comprehensive and multidimensional approach to disease assessment.

Patient-reported outcomes are an essential part of this framework and play a critical role in clinical decision making by providing insights into the patient’s experience and helping families and care teams establish treatment goals. Questionnaires such as PROMIS, PedQOL (Pediatric Quality of Life Inventory), and CHAQ have been widely used in pediatric chronic diseases like JIA and SLE to monitor disease burden and treatment effectiveness.^7,16^

This study provided evidence that the T-scores of the PROMIS Pediatric measures of pain interference, pain behavior, and mobility had strong association with self-reported disease severity and sensitivity to change following effective treatment. These findings suggest that these HRQOL domains may be robust measures for assessing disease status in patients with CNO. Interestingly, these same variables have also shown strong performance in JIA subgroups, further supporting their relevance in pediatric rheumatic disease.^7^ In contrast, T-scores associated with PROMIS Pediatric measures of physical activity, fatigue, strength impact, and upper extremity variables did not show consistent significance across all the comparisons with external measures or over time. Specifically, physical activity T-scores did not show significant change over time, a finding consistent in studies of JIA and cSLE.^7^ This suggests that the current construct of physical activity measurement within PROs may not be sufficiently sensitive to detect changes in children with rheumatic diseases. Alternatively, patients may not experience improved physical activity within the observed period despite disease improvement.

A previous small cohort study from our group suggested minor improvement in mobility and pain T-scores following treatment, but these findings did not reach statistical significance due to the limited sample size. ^17^ By systematically examining the performance of PROMIS Pediatric measures in a larger and more diverse patient population, this current study provides more generalizable insights into the utility of these measures in CNO.

Our data shows significant inter-individual variation in PROMIS T-scores, particularly within the inactive disease group. There was notable overlap in T-scores across different disease severity categories, emphasizing the inherent variability in patient-reported measurements. Unlike patients with JIA, who often present with persistent symptoms, CNO symptoms may be more intermittent, fluctuating from day to day^18^. As PROMIS Pediatric measures assess symptoms over the previous seven days, responses may be influenced by short-term symptom variability rather than long-term disease activity. PROMIS T-scores should be interpreted cautiously in conjunction with comprehensive clinical assessment. Further studies are needed to assess the test-retest reliability and daily variation of these symptoms and associated measures.

While findings support the use of PROMIS Pediatric measures in CNO, there are several considerations that warrant further investigation. One limitation of our study is that we did not compare PROMIS Pediatric T-scores to MRI findings, which could provide an additional understanding of disease status. Indeed, clinically silent lesions of the vertebral column may be missed using the PROMIS tool but can cause significant complications and would warrant treatment escalation^19^. Additionally, we did not assess the burden of questionnaire completion on families, which is an important factor in determining feasibility for routine clinical use. As discussed above, another key consideration is the variability in individual responses. We did not perform intra-individual variation testing of the PROMIS instruments, and our sample size for second-line treatment outcomes was relatively small. Some of our cohorts and comparison groups had small sample size that gives the results less power. The study would benefit from additional collaboration with international groups studying CNO. While our study highlights the value of PROMIS Pediatric measures in tracking disease burden, its variability at the individual level suggests that it should not be used in isolation in routine clinical practice. The combination of patient-reported data and clinical examination (including scans such as MRI) provides a comprehensive solution.

## Data Availability

All data produced in the present study are available upon reasonable request to the authors

## Conclusion

This study provides evidence for the PROMIS Pediatric measures of mobility, pain behavior, pain interference to be used in clinical research setting and potential disease monitoring for CNO with good performance of convergence, divergence and sensitivity to change in the English spoken patients from a large international patient registry.

