## Supplemental Table 1 for "Validation of Patient-Reported Outcomes Measurement Information System^®^ (PROMIS^®^) Pediatric Measures for Children with Chronic Nonbacterial Osteomyelitis"

***Supplementary Material***

**<u>Supplementary Table 1. PROMIS Pediatric and CHAQ Score Changes When Patients Reported Better or unchanged Comparing to those Reported Worse</u>**

| Variables | Predictors | Mean | SD | P-Value | Adjusted p value (update) |
|---|---|---|---|---|---|
| Fatigue n=196 | Better (n=76) | -1.9 | 12 | 0.02 | 0.04 |
| | Unchanged (n=92) | -0.2 | 11 | 0.08 | 0.10 |
| | Worse (n=28) | 4.3 | 13 | NA | NA |
| Pain Behavior n=198 | Better (n=78) | -4 | 11 | <0.001 | <0.001 |
| | Unchanged (n=91) | -1.9 | 9 | <0.001 | <0.001 |
| | Worse (n=29) | 9.8 | 15 | NA | NA |
| Pain Interference n=198 | Better (n=78) | -0.5 | 11 | 0.01 | 0.02 |
| | Unchanged (n=92) | -1.5 | 8 | 0.003 | 0.02 |
| | Worse (n=28) | 4.7 | 12 | NA | NA |
| CHAQ n=94 | Better (n=47) | -0.2 | 1 | 0.03 | 0.04 |
| | Unchanged (n=38) | 0 | 0 | 0.35 | 0.40 |
| | Worse (n=9) | 0.2 | 1 | NA | NA |
| Mobility n=198 | Better (n=78) | 0.8 | 7 | 0.02 | 0.04 |
| | Unchanged (n=92) | 1.7 | 6 | 0.004 | 0.02 |
| | Worse (n=28) | -3.1 | 11 | NA | NA |
| Upper Extremity n=198 | Better (n=78) | 1.1 | 5 | 0.06 | 0.08 |

| | | | | | |
|---|---|---|---|---|---|
| | Unchanged (n=92) | 1.4 | 5 | 0.03 | 0.04 |
| | Worse (n=28) | -1.0 | 4 | NA | NA |
| Physical Activity n=200 | Better (n=79) | 1.8 | 9 | 0.01 | 0.02 |
| | Unchanged (n=92) | 1.5 | 9 | 0.01 | 0.02 |
| | Worse (n=29) | -3.7 | 10 | NA | NA |
| Strength Impact n=197 | Better (n=77) | 1.9 | 11 | 0.65 | 0.69 |
| | Unchanged (n=92) | 1.2 | 12 | 0.85 | 0.85 |
| | Worse (n=28) | 0.8 | 8 | NA | NA |
